# Differential Impact of Smoking on Intracerebral Hemorrhage Based on Cerebral Microbleed Status: A Case-Control Study

**DOI:** 10.1101/2025.08.14.25333729

**Authors:** Takehito Kuroda, Tomotaka Tanaka, Satoshi Saito, Sonu Bhaskar, Hiroyuki Ishiyama, Ryoma Inui, Yuma Shiomi, Yuriko Nakaoku, Soshiro Ogata, Yoshiaki Morita, Etsuko Ozaki, Nagato Kuriyama, Kunihiro Nishimura, Masatoshi Koga, Jin Nakahara, Kazunori Toyoda, Masafumi Ihara

## Abstract

**Background:** While smoking is a major cause of cardiovascular diseases, its association with intracerebral hemorrhage (ICH) remains controversial. Cerebral microbleeds (CMBs) serve as an imaging indicator of ICH, yet ICH can occur independently of CMBs, suggesting distinct underlying mechanisms. We investigated the impact of smoking on ICH after stratifying individuals by CMB presence or absence.

**Methods:** We conducted a retrospective case-control study and enrolled patients with ICH as their first stroke and controls with non-stroke neurological conditions admitted to our center between 2017 and 2021. The participants were classified by CMB presence on magnetic resonance imaging and smoking status (current/past/never). We performed multivariable logistic regression analysis to assess the association between smoking and ICH, stratified by CMB status. We also assessed whether smoking cessation was linked to a lower ICH risk than current smoking, specifically among past smokers stratified by the duration of cessation.

**Results:** We analyzed 487 patients with ICH (median age 70.0 years, interquartile range[IQR] [58.0–81.0], 41.5% female, 53.6% with CMBs) and 322 controls (median age 71.0 years, IQR [58.8–79.0], 47.5% female, 13.4% with CMBs). In the individuals without CMBs, current smoking was more frequently observed in patients with ICH than in controls (25.7% vs. 14.0%, p<0.001) and was independently associated with ICH following adjustment for potential confounders (adjusted odds ratio[aOR] 1.84, 95% confidence interval[CI] 1.02–3.31, p=0.042). Long-term smoking cessation (>10 years) was associated with a lower ICH risk than current smoking (aOR 0.31, 95%CI 0.14–0.67, p=0.003), while short-term cessation (≤10 years) showed no significant difference. The individuals with CMBs demonstrated no significant association between smoking status and ICH.

**Conclusions:** Current smoking is significantly associated with ICH in individuals without CMBs but not in individuals with CMBs. Long-term smoking cessation might mitigate ICH risk in individuals without CMBs, emphasizing early smoking cessation as a preventive strategy. Assessing smoking history may aid in identifying those at ICH risk.

## Introduction

Intracerebral hemorrhage (ICH) affects 3.41 million people annually worldwide, representing 27.9% of all strokes according to the Global Burden of Diseases, Injuries, and Risk Factors Study 2019,^1^ and is associated with high rates of severe disability and mortality.^2,3^ Although substantial advancements in acute reperfusion therapy have improved outcomes for patients with acute ischemic stroke (AIS) over the past two decades, there remains no definitive curative treatment for ICH.^3^ Therefore, primary prevention strategies are critical for reducing the incidence of ICH.

Smoking increases cardiovascular risk.^4^ While its association with ischemic stroke has been consistently demonstrated,^5^ the relationship between smoking and ICH remains unclear. Some studies have reported that smoking is a significant risk factor for ICH,^6,7^ particularly in younger individuals,^8,9^ whereas others have failed to identify a significant association.^10,11^ Understanding the impact of smoking cessation is also essential for better evaluating smoking as a potentially modifiable risk factor. However, evidence regarding the benefits of smoking cessation for ICH prevention is limited. For instance, a cohort study reported no statistically significant reduction in ICH risk among past smokers compared to current smokers (hazard ratio [HR] 0.82, 95% confidence interval [CI]: 0.64–1.06).^12^ These discrepancies may be attributed to the variation in ICH etiology across various population groups, particularly between younger and older individuals.

Cerebral microbleeds (CMBs) are a valuable imaging biomarker detected on magnetic resonance imaging (MRI) as hypointense foci, particularly on gradient-echo T2 star-weighted imaging (GRE T2*WI).^13–15^ The presence of CMBs reflects prior blood extravasation and bleeding-prone microangiopathy.^16^ CMBs, which are an independent risk factor for ICH,^17,18^ are found in 19% of patients with AIS undergoing reperfusion therapy, significantly increasing the odds of symptomatic ICH,^19^ hemorrhagic transformation, poor functional outcomes, and mortality at 90 days.^20^ A systematic review reported that CMBs are observed in approximately 60% of patients with ICH compared with 5% of healthy participants (mean age 52.9–64.4 years).^21^ A longitudinal study further demonstrated a significantly higher HR (50.2) for developing ICH among individuals with CMBs at baseline than among those without CMBs over a mean follow-up period of 3.6 years.^22^ CMBs are also associated with cardiovascular risk factors, such as hypertension, diabetes, and smoking, as well as age and cerebral amyloid angiopathy (CAA).^17,23,24^ Consequently, ICH with CMBs is characterized by vascular fragility, which refers to the structural and functional vulnerability of blood vessels caused by a complex interplay of microangiopathy, amyloid deposition, and chronic inflammation. This multifactorial nature may obscure the specific contribution of smoking to ICH risk. In contrast, ICH without CMBs is less likely to be associated with CAA, and, correspondingly, shows a more specific association with microangiopathy than ICH with CMBs. This distinction may allow for a more precise assessment of the effects of smoking on ICH.

However, previous studies have not examined whether CMB status influences the relationship between ICH and smoking. Since CMBs represent different underlying pathologies, stratifying by CMB presence may clarify the inconsistent findings in the literature and identify specific populations at higher smoking-related ICH risk. Thus, we hypothesized that smoking may independently influence ICH risk in individuals without CMBs. To clarify our hypothesis, we examined the impact of smoking on ICH after stratifying individuals by CMB presence or absence.

## Methods

### Study Design, Setting, and Period

We conducted a case-control study to investigate the associations between ICH and smoking at the Department of Cerebrovascular Medicine and Neurology at the National Cerebral and Cardiovascular Center (NCVC), a major stroke center in Japan. Patients admitted between January 2017 and December 2021 were included.

### Study Participants

The case group included consecutive patients with ICH and no prior history of stroke. The control group consisted of patients with non-stroke neurological diseases and no history of stroke. Patient data were derived from the NCVC Stroke Registry (ClinicalTrials.gov Identifier: NCT02251665), which prospectively registered patients with stroke and other neurological diseases.^25^ Patients in both groups were excluded if smoking status was unavailable or if GRE T2*WI imaging for CMBs was not performed during hospitalization.

### Clinical Data Collection

We reviewed patient electronic medical records to extract data on age, sex, premorbid modified Rankin Scale (mRS) score, body mass index (BMI), medical history including hypertension (systolic/diastolic blood pressure ≥140/90 mmHg or antihypertensive medication use), diabetes mellitus (glycated hemoglobin level ≥6.5% and blood glucose ≥126 mg/dL or antidiabetic medication or insulin use), dyslipidemia (low-density lipoprotein cholesterol ≥140 mg/dL, high-density lipoprotein cholesterol <40 mg/dL, or antihyperlipidemic drug use), and antithrombotic drug use, smoking status, and current drinking status. BMI was calculated as weight in kilograms divided by height in meters squared, with BMI <18.5 kg/m² defined as underweight. Current drinking was defined as consuming alcohol at least 3 days per week, with a minimum daily intake of one serving. Smoking status was classified as follows^26^:

1. Current smoking: smoking ≥100 cigarettes in their lifetime and currently smoking,
2. Past smoking: smoking ≥100 cigarettes in their lifetime but not currently smoking, and
3. Never smoking: no smoking history or smoking <100 cigarettes in their lifetime.

As additional smoking-related information, the duration of smoking cessation in past smoking was also recorded when available. Among heavy smokers (i.e., ≥20 pack-years), the Framingham Heart Study demonstrated that the cardiovascular risk difference between past and never smokers was no longer significant after 10 years of smoking cessation. Moreover, a previous review reported that smoking cessation for >10 years was associated with a 2–4-fold difference in stroke risk compared with current smoking.^5,27^ On the basis of these findings, the smoking cessation duration was categorized as short-term (≤10 years) or long-term (>10 years).

### CMB Evaluation

CMBs were evaluated using GRE T2*WI, according to the STandards for ReportIng Vascular changes on nEuroimaging (STRIVE), defined as small areas (generally 2–5 mm in diameter, but sometimes up to 10 mm) of signal void with associated blooming.^28^

The sequence parameters of GRE T2*WI were as follows: slice thickness, 4.0 mm; interslice gap, 2.0 mm; echo time, 12 ms; repetition time, 400–500 ms; and flip angle, 20°. MRI was performed using either a 1.5-Tesla scanner (Magnetom Sonata or Vision; Siemens Medical Solutions, Erlangen, Germany) or 3.0-Tesla scanner (Magnetom Verio or Spectra; Siemens Medical Solutions, Erlangen, Germany).

All patients with ICH and controls were subsequently categorized according to CMB presence on MRI: patients with ICH without and with CMBs (ICH/CMB(−) and ICH/CMB(+), respectively) and controls without and with CMBs (Control/CMB(−) and Control/CMB(+), respectively) (Figure 1). For analysis, we defined two main comparison groups: the CMB(–) groups, comparisons between the ICH/CMB(−) and the Control/CMB(−) groups; and the CMB(+) groups, comparisons between the ICH/CMB(+) and the Control/CMB(+) groups.

**Figure 1.**
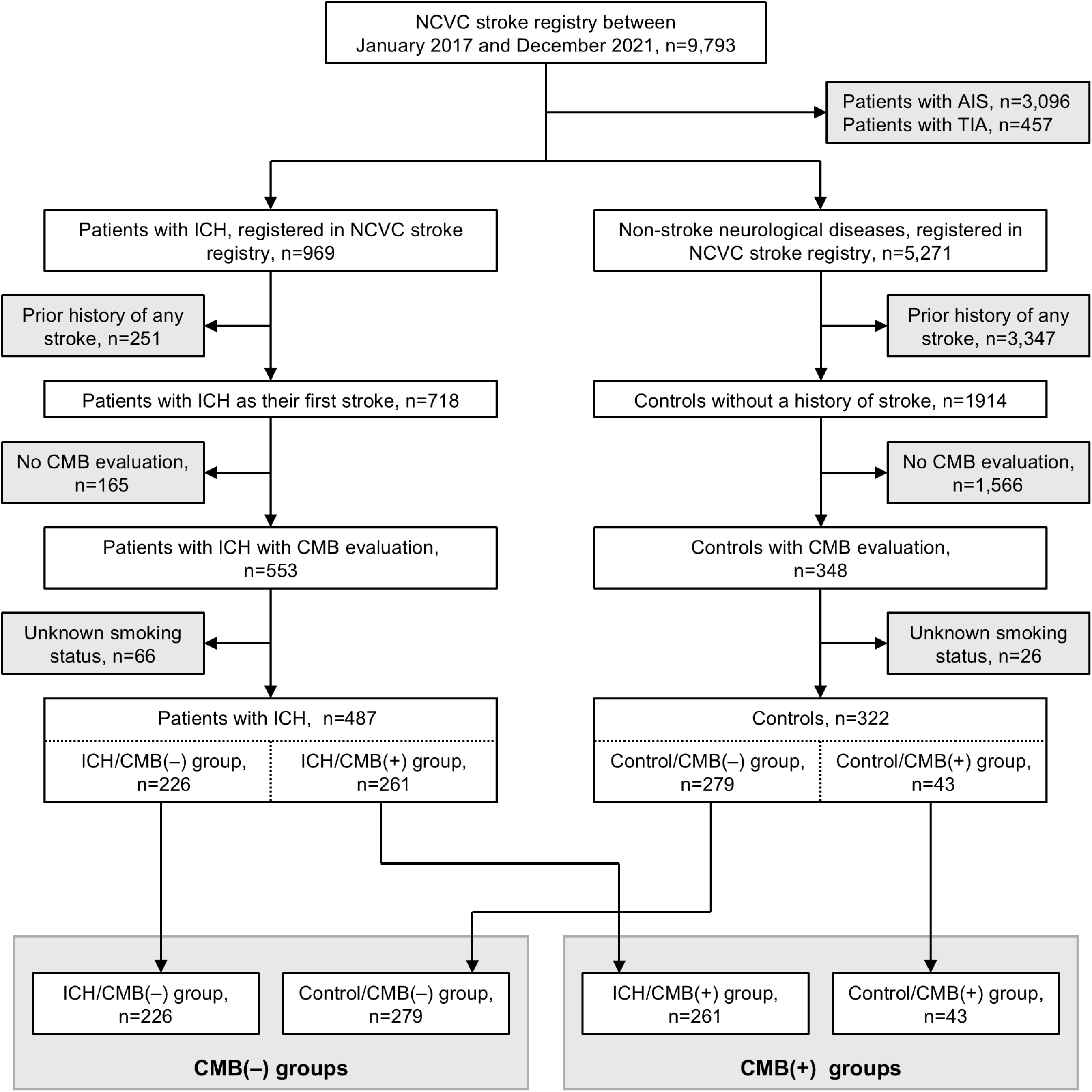
Study flowchart. From the NCVC stroke registry between January 2017 and December 2021, we included patients with ICH and individuals with non-stroke neurological disease as controls. After excluding cases of prior stroke and missing CMB evaluation, 487 patients with ICH and 322 controls with CMB evaluation were stratified into the CMB(+) and CMB(–) groups for comparative analysis. Abbreviations: ICH, intracerebral hemorrhage; NCVC, National Cerebral and Cardiovascular Center; CMB, cerebral microbleed.

### Statistical Analysis

In all analyses, we evaluated all individuals and performed separate analyses for the CMB(–) and the CMB(+) groups. We compared the characteristics between patients with ICH and controls using the chi-square test for categorical variables and the Mann–Whitney U test for continuous variables. Continuous variables are presented as median (interquartile range [IQR]), whereas categorical variables are presented as number (%). For patients with ICH, we compared hematoma volumes, etiologies, and locations between the ICH/CMB(+) and the ICH/CMB(−) groups. To assess the independent association of current smoking with ICH risk, we performed multivariable logistic regression analysis and calculated odds ratios (ORs). We constructed two models: Model 1 was adjusted for basic demographic factors (age and sex), whereas Model 2 was additionally adjusted for premorbid mRS score and modifiable risk factors for ICH identified in previous studies (hypertension, underweight, and current drinking status).^29–31^ The adjusted ORs (aORs) with 95% CIs were calculated.

Furthermore, we investigated whether the impact of smoking on ICH varied with smoking cessation periods (current smoking versus [vs.] short-term and long-term cessation) with multivariate logistic regression analyses adjusted by the factors in Model 2. We also conducted sensitivity analyses with multivariate logistic regression (Model 2) to evaluate potential effect modification by age and hypertension history, both major risk factors for ICH. Patient age was dichotomized into ≥65 years and < 65 years, since individuals aged 65 and older are generally defined as elderly in Japan. Missing data were handled with pairwise deletion. Statistical analyses were conducted using SPSS Statistics for Windows, version 29.0 (IBM Corp., Armonk, NY, USA), with p<0.05 considered statistically significant.

### Ethical Considerations

Ethical approval was obtained from the local institutional review board (M23-073-13), which waived the requirement for written informed consent because clinical information obtained in routine clinical practice was used, no additional invasive procedures or costs were imposed on the patients, and the information was sufficiently anonymized. All procedures were conducted in accordance with the principles of the Declaration of Helsinki. This study was also reported in accordance with the Strengthening the Reporting of Observational Studies in Epidemiology (STROBE) guidelines for case-control studies. A completed STROBE checklist is available as Supplemental Material.

## Results

### Participant Selection

We identified 969 patients with ICH and 5,271 patients with non-stroke neurological diseases from the NCVC Stroke Registry (January 2017 to December 2021), which includes 9,793 patients. After excluding individuals with a stroke history, missing CMB evaluation, or unknown smoking status, the final analysis comprised 487 patients with ICH and 322 controls (Figure 1). The controls included patients with various diseases/disorders, such as benign paroxysmal positional vertigo, epilepsy, carotid artery stenosis/occlusion, and other diseases (Table S1).

### Baseline Characteristics

#### Comparing all patients with ICH to all controls

The median ages among patients with ICH and controls were 70.0 years (IQR 58.0–81.0) and 71.0 years (IQR 58.8–79.0) (p=0.733), respectively. Most of the patients with ICH and controls were male (58.5% vs. 52.5%, p=0.090, respectively) who were of normal weight or higher (82.1% vs. 85.4%, p=0.254, respectively). A similar proportion of the patients with ICH and controls had diabetes mellitus (22.8% vs. 18.6%, p=0.295) or dyslipidemia (28.1% vs. 34.5%, p=0.056), were on antithrombotics (17.5% vs. 23.0%, p=0.053), or were past smokers (20.9% vs. 24.5%, p=0.230) or never smokers (57.7% vs. 61.8%, p=0.245). CMB prevalence was significantly higher among the patients with ICH than in controls (53.6% [261 patients] vs. 13.4% [43 patients] p<0.001) (Table 1). There was a significantly higher proportion of the patients with ICH who had hypertension (89.5% vs. 54.3%, p<0.001), were current smokers (21.4% vs. 13.7%, p=0.006), were current drinkers (42.5% vs. 25.3%, p<0.001), and had CMBs (53.6% vs. 13.4%, p<0.001), when compared with the controls. The patients with ICH had a significantly better premorbid mRS score than controls (median 0 IQR [0–0] vs. 0 [0–1], p=0.035) (Table 1).

**Table 1.**
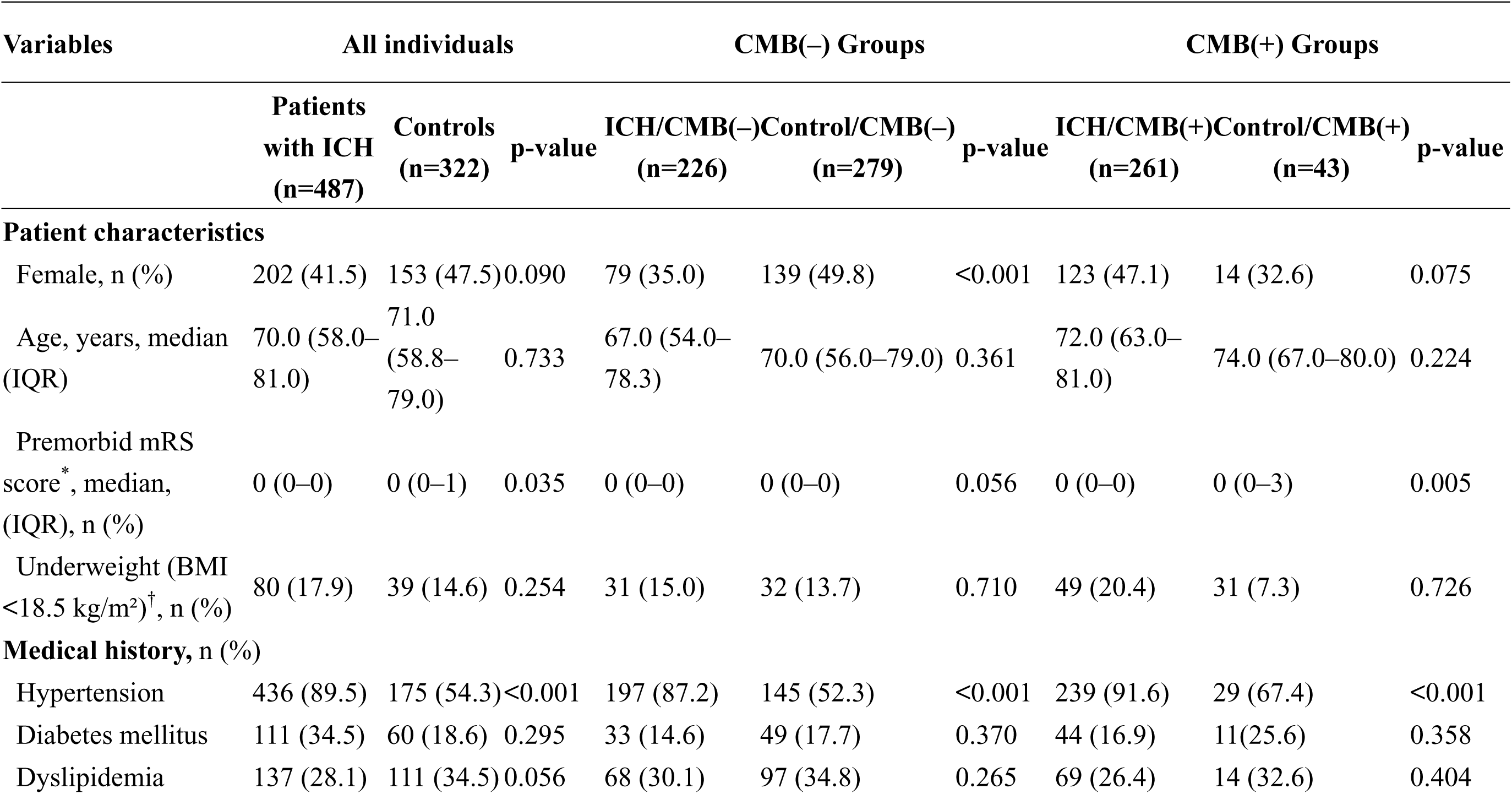
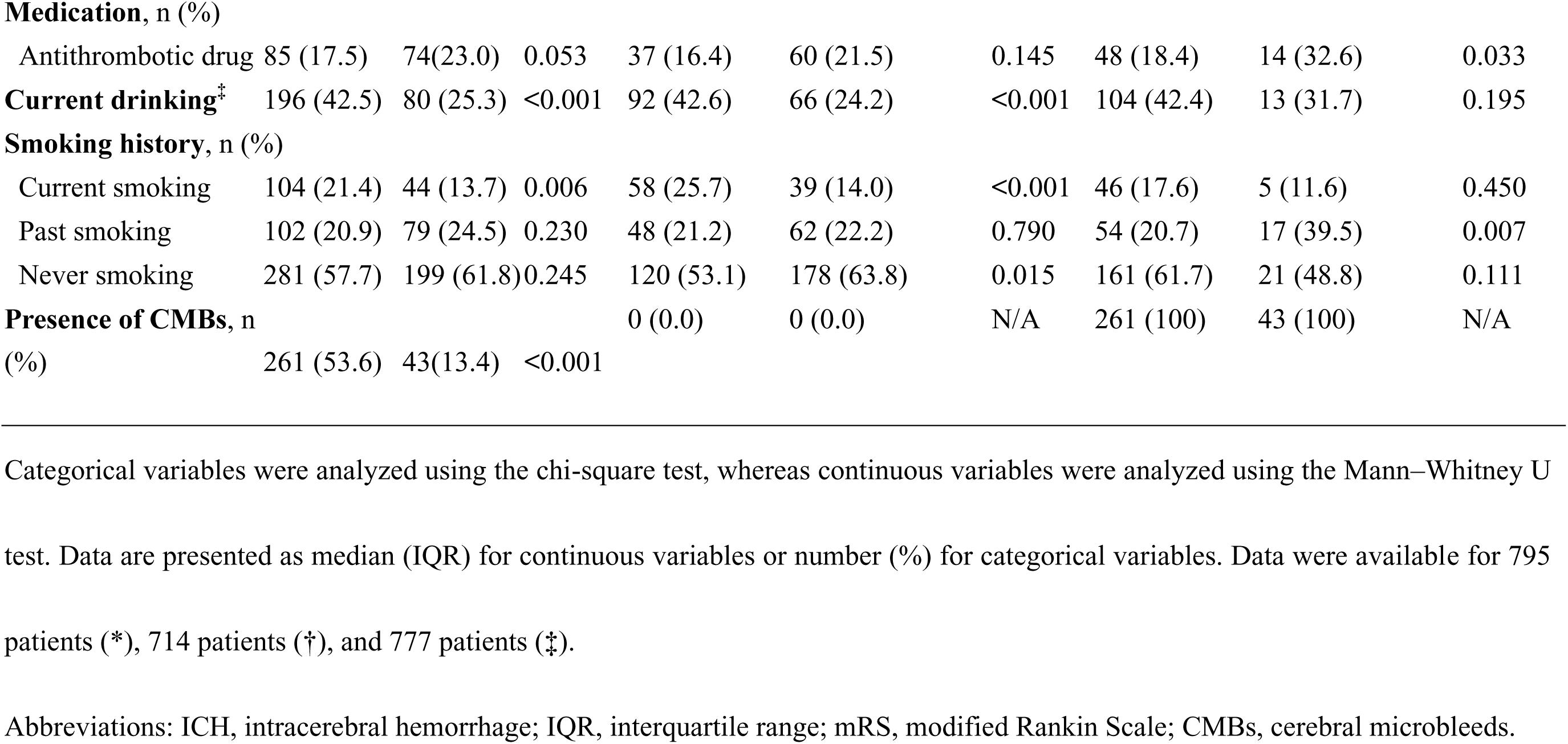
Baseline characteristics of patients with intracerebral hemorrhage and controls stratified by cerebral microbleed status.

| Variables | All individuals |  |  | CMB(-) Groups |  |  | CMB(+) Groups |  |  |
| --- | --- | --- | --- | --- | --- | --- | --- | --- | --- |
|  | Patients<br>with ICH<br>(n=487) | Controls<br>(n=322) | p-value | ICH/CMB(-)<br>(n=226) | Control/CMB(-)<br>(n=279) | p-value | ICH/CMB(+)<br>(n=261) | Control/CMB(+)<br>(n=43) | p-value |
| **Patient characteristics** |  |  |  |  |  |  |  |  |  |
| Female, n (%) | 202 (41.5) | 153 (47.5) | 0.090 | 79 (35.0) | 139 (49.8) | <0.001 | 123 (47.1) | 14 (32.6) | 0.075 |
| Age, years, median<br>(IQR) | 70.0 (58.0–<br>81.0) | 71.0<br>(58.8–<br>79.0) | 0.733 | 67.0 (54.0–<br>78.3) | 70.0 (56.0–79.0) | 0.361 | 72.0 (63.0–<br>81.0) | 74.0 (67.0–80.0) | 0.224 |
| Premorbid mRS<br>score<sup>*</sup>, median,<br>(IQR), n (%) | 0 (0–0) | 0 (0–1) | 0.035 | 0 (0–0) | 0 (0–0) | 0.056 | 0 (0–0) | 0 (0–3) | 0.005 |
| Underweight (BMI<br><18.5 kg/m<sup>2</sup>)<sup>†</sup>, n (%) | 80 (17.9) | 39 (14.6) | 0.254 | 31 (15.0) | 32 (13.7) | 0.710 | 49 (20.4) | 31 (7.3) | 0.726 |
| **Medical history, n (%)** |  |  |  |  |  |  |  |  |  |
| Hypertension | 436 (89.5) | 175 (54.3) | <0.001 | 197 (87.2) | 145 (52.3) | <0.001 | 239 (91.6) | 29 (67.4) | <0.001 |
| Diabetes mellitus | 111 (34.5) | 60 (18.6) | 0.295 | 33 (14.6) | 49 (17.7) | 0.370 | 44 (16.9) | 11(25.6) | 0.358 |
| Dyslipidemia | 137 (28.1) | 111 (34.5) | 0.056 | 68 (30.1) | 97 (34.8) | 0.265 | 69 (26.4) | 14 (32.6) | 0.404 |
| **Medication, n (%)** |  |  |  |  |  |  |  |  |  |
| Antithrombotic drug | 85 (17.5) | 74(23.0) | 0.053 | 37 (16.4) | 60 (21.5) | 0.145 | 48 (18.4) | 14 (32.6) | 0.033 |
| **Current drinking<sup>‡</sup>** | 196 (42.5) | 80 (25.3) | <0.001 | 92 (42.6) | 66 (24.2) | <0.001 | 104 (42.4) | 13 (31.7) | 0.195 |
| **Smoking history, n (%)** |  |  |  |  |  |  |  |  |  |
| Current smoking | 104 (21.4) | 44 (13.7) | 0.006 | 58 (25.7) | 39 (14.0) | <0.001 | 46 (17.6) | 5 (11.6) | 0.450 |
| Past smoking | 102 (20.9) | 79 (24.5) | 0.230 | 48 (21.2) | 62 (22.2) | 0.790 | 54 (20.7) | 17 (39.5) | 0.007 |
| Never smoking | 281 (57.7) | 199 (61.8) | 0.245 | 120 (53.1) | 178 (63.8) | 0.015 | 161 (61.7) | 21 (48.8) | 0.111 |
| **Presence of CMBs, n** |  |  |  | 0 (0.0) | 0 (0.0) | N/A | 261 (100) | 43 (100) | N/A |
| (%) | 261 (53.6) | 43(13.4) | <0.001 |  |  |  |  |  |  |

Categorical variables were analyzed using the chi-square test, whereas continuous variables were analyzed using the Mann–Whitney U
test. Data are presented as median (IQR) for continuous variables or number (%) for categorical variables. Data were available for 795
patients (\*), 714 patients (†), and 777 patients (‡).
Abbreviations: ICH, intracerebral hemorrhage; IQR, interquartile range; mRS, modified Rankin Scale; CMBs, cerebral microbleeds.

#### Comparing the ICH/CMB(+) group to the ICH/CMB(−) group

The ICH/CMB(+) group was older (72.0 vs. 67.0 years, p<0.001) and more likely to be female (47.1% vs. 35.0%, p=0.007), have CAA (14.2% vs. 0.4%, p<0.001), and have thalamic hemorrhages (34.1% vs. 23.5%, p=0.010). In the ICH/CMB(−) group, only one patient was diagnosed with CAA, who presented with cortical siderosis. The hematoma volumes were similar between the ICH/CMB(−) and ICH/CMB(+) groups (median 9.6 IQR [4.2–25.5] mL vs. 10.1 [4.6–23.4] mL, p=0.473) (Table S2).

#### Comparing the ICH/CMB(−) group to the Control/CMB(−) group

There was a significantly higher proportion of males in the ICH/CMB(−) group (65.0% vs. 50.2%, p<0.001), who were current drinkers (42.6% vs. 24.2%, p<0.001) and current smokers (25.7% vs. 14.0%, p<0.001), with hypertension (87.2% vs. 52.3%, p<0.001) when compared with the Control/CMB(−) group (Table 1). Conversely, there was a significantly lower proportion of never smokers among the ICH/CMB(−) group when compared with the Control/CMB(−) group (53.1% vs. 63.8%, p=0.015) (Table 1).

#### Comparing the ICH/CMB(+) group to the Control/CMB(+) group

There was a significantly higher proportion in the ICH/CMB(+) group with hypertension (91.6% vs. 67.4%, p<0.001) than in the Control/CMB(+) group. The ICH/CMB(+) group was less likely to be on antithrombotic drug use (18.4% vs. 32.6%, p=0.033) and less likely to be past smokers (20.7% vs 39.5%, p=0.007) when compared with the Control/CMB(+) group. The ICH/CMB(+) group had significantly better premorbid mRS scores than the Control/CMB(+) group (median 0 [IQR 0–0] vs. 0 [0–3], p=0.005) (Table 1).

#### Impact of Smoking on ICH

Multivariable logistic regression analyses demonstrated that current smoking was significantly associated with increased odds of ICH in all individuals after adjustment for Model 2 covariates (aOR 1.71, 95% CI 1.03–2.84, p=0.039) (Table 2). This association was particularly pronounced in the CMB(–) group (Crude: 2.11 [95% CI, 1.35–3.34]; Model 1: 1.95 [1.21–3.14]; Model 2: 1.84 [1.02–3.31]). No significant association of current smoking with ICH was observed in the CMB(+) group.

**Table 2.** Impact of current smoking on ICH.

| Model | All individuals |  | CMB(−) |  | CMB(+) |  |
| --- | --- | --- | --- | --- | --- | --- |
|  | aOR (95% CI) | p-value | aOR (95% CI) | p-value | aOR (95% CI) | p-value |
| Crude | 1.72<br>(1.17–2.52) | 0.006 | 2.11<br>(1.35–3.34) | 0.001 | 1.63<br>(0.61–4.34) | 0.334 |
| Model 1 | 1.82<br>(1.21–2.73) | 0.004 | 1.95<br>(1.21–3.14) | 0.006 | 1.70<br>(0.61–4.75) | 0.309 |
| Model 2 | 1.71<br>(1.03–2.84) | 0.039 | 1.84<br>(1.02–3.31) | 0.042 | 1.41<br>(0.41–4.82) | 0.586 |

Data are presented as aOR with 95% CI. Model 1 was adjusted for age and sex. Model 2 was additionally adjusted for premorbid modified Rankin Scale score, hypertension, underweight (body mass index <18.5 kg/m<sup>2</sup>), and current drinking status.
Abbreviations: ICH, intracerebral hemorrhage; CMB, cerebral microbleed; aOR, adjusted odds ratio; CI, confidence interval.

#### Effect of Smoking Cessation Period

The median smoking cessation period among past smokers was significantly shorter in the patients with ICH than in the controls (median 12.0 IQR, [6.0–7.0] years vs. 20.0 [12.0–38.0] years, p=0.018). This difference was also observed in the CMB(–) group (the ICH/CMB(−) group: 12.0 [4.0–20.0] years vs. the Control/CMB(−) group: 20.0 [10.0–38.0] years, p=0.003). However, in the CMB(+) group, the cessation periods were comparable between patients with ICH and controls (the ICH/CMB(+) group: 19.0 [7.5–37.0] years vs. the Control/CMB(+) group: 16.0 [10.0–36.0] years, p=0.716).

The multivariate logistic regression analysis showed that long-term smoking cessation was significantly associated with reduced odds of ICH in all individuals (aOR 0.31, 95% CI 0.16– 0.60, p<0.001) and in the CMB(–) group (aOR 0.31, 95% CI 0.14–0.67, p=0.003) (Figure 2).

**Figure 2.**
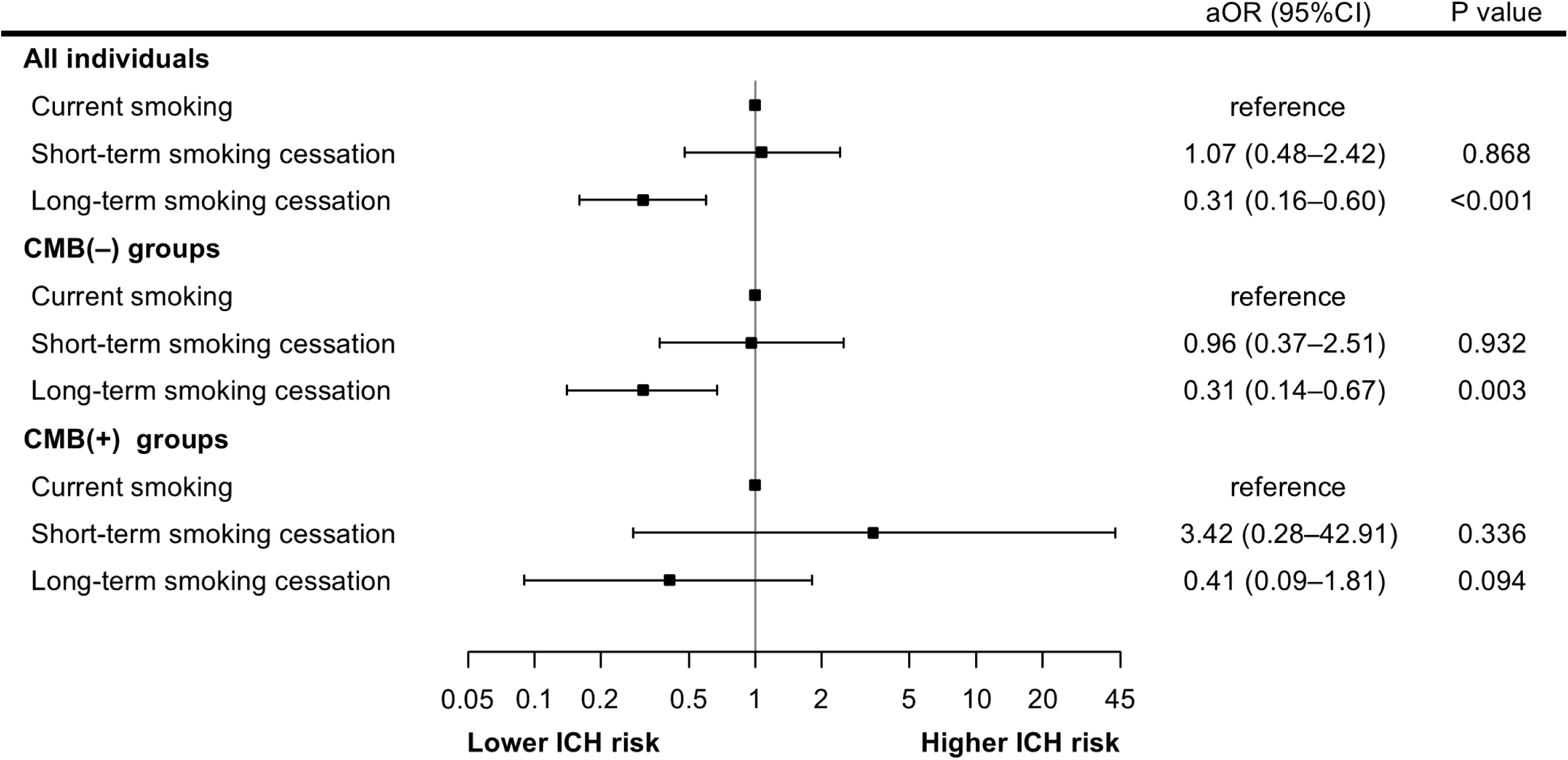
Association of ICH with smoking cessation period according to CMB status. Forest plots showing aOR for the association between ICH and smoking cessation period (short-term: ≤10 years; long-term: ≥11 years), compared with current smoking. All analyses were adjusted for age, sex, premorbid mRS score, and modifiable risk factors (hypertension, underweight, and current drinking status). The analysis included 309 available patients with current or past smoking status. Abbreviations: ICH, intracerebral hemorrhage; CMB, cerebral microbleed; aOR, adjusted odds ratio; CI, confidence interval; mRS, modified Rankin Scale.

In contrast, short-term cessation showed no significant association with ICH across any groups. In the CMB(+) group, neither short-term nor long-term smoking cessation was significantly associated with ICH (Figure 2).

#### Sensitivity analyses

In age-stratified analyses using a cutoff of 65 years (Table S3), current smoking was not associated with ICH in any subgroup, regardless of age or CMB status. When stratified by hypertension status (Table S4), current smoking showed a trend toward an association with ICH among all individuals with hypertension (aOR 1.77, 95% CI 0.94–3.13, p=0.077) and reached statistical significance in the CMB(–) group with hypertension (aOR 2.03, 95% CI 1.001–4.12, p=0.050). No significant associations were observed among individuals without hypertension in the CMB(–) and CMB(+) groups.

## Discussion

This study investigated the impact of smoking on ICH risk after stratifying individuals by CMB presence or absence. Current smoking was significantly associated with increased ICH risk in the CMB(–) group but not in the CMB(+) group. Moreover, long-term smoking cessation of more than 10 years appeared to mitigate this risk, particularly in the CMB(–) group, while short-term cessation showed no significant benefit. These findings suggest that the impact of smoking on ICH may differ depending on the underlying vascular pathology.

Several mechanisms have been proposed to explain how smoking increases ICH risk. For instance, smoking rapidly induces transient increases in blood pressure,^32^ which can exacerbate the risk of hemorrhage in already vulnerable vessels.^33^ Additionally, smoking contributes to structural changes in the arterial walls, including aneurysm formation and blood-brain barrier (BBB) damage.^34,35^ Tobacco smoke contains abundant reactive oxygen species (ROS), which induce oxidative damage to vessel walls, leading to increased vascular fragility.^36^ Moreover, smoking impairs BBB integrity through the upregulation of proinflammatory cytokines including tumor necrosis factor-alpha (TNF-α), and increases nitric oxide and ROS production, all of which weaken endothelial barrier function.^37^ These smoking-mediated alterations collectively increase susceptibility to ICH.

In the present study, these effects on ICH risk were primarily evident in the CMB(–) group. The ICH/CMB(−) group was characterized by male predominance, higher hypertension rates, and increased current drinking compared with the Control/CMB(−) group. The accumulation of these characteristics increased their susceptibility to vascular injury and may contribute to the more apparent association with smoking. Additionally, the underlying pathology is less likely to be CAA and more likely to be microangiopathy, with vascular structural changes and remodeling that are less pronounced than those in the CMB(+) group.^38^ Currently, hypertensive ICH is thought to be caused by a lipohyalinotic change, a combination of hyalinization and lipid deposition in the vessel wall, rather than microaneurysms (Charcot– Bouchard aneurysms [CBA]) that were traditionally considered the source of bleeding.^39^ However, the roles of aneurysms of small vessels in ICH remain contested. A recent pathological study examining 2,749 autopsy cases (mean age 80.9±8.8 years) found CBAs in only 12 cases.^40^ However, in early-stage hypertension, vessels may retain enough flexibility to form aneurysms, whereas in advanced stages, vessel wall thickening and fibrosis might instead prevent microaneurysm formation. Early radiographic studies using barium sulfate contrast observed CBAs in 46% of patients with hypertension, with the highest prevalence (71%) in patients aged 65–69 years, followed by 60% in patients aged 70–74 years and 47% in patients aged >75 years.^41^ Smoking is also implicated in aneurysm formation in subarachnoid hemorrhage (SAH), primarily through mechanisms that compromise vessel wall integrity and lead to endothelial dysfunction and chronic inflammation.^42–44^ Therefore, in the CMB(−) group where hypertensive vasculopathy predominates and CAA is rare, smoking effects might be more directly manifested through mechanisms similar to those observed in aneurysm formation and SAH.

In contrast to the CMB(−) group, the CMB(+) group showed no significant association between current smoking and ICH. The baseline characteristics revealed important differences, with the Control/CMB(+) group showing higher antithrombotic drug use, greater prevalence of past smoking, and worse premorbid functional status compared with patients with ICH. This suggests that controls represented individuals with substantial cardiovascular risk profiles despite no stroke history, creating baseline imbalances that may have reduced the ability to detect smoking-specific effects. Furthermore, the underlying pathology appears to be more complex. It is characterized by a more multifactorial vascular fragility, including contributions from CAA and other age-related and inflammatory changes, which may obscure the specific effects of smoking. Approximately 14% of cases in the ICH/CMB(+) group were attributed to CAA, although hypertension accounted for the majority, whereas CAA was rarely observed in the ICH/CMB(−) group. A previous report also showed that hypertensive vasculopathy remains a primary mechanism and that CAA is a significant contributor to ICH, which is caused by vascular fragility due to amyloid deposition.^45^ As CAA development is primarily influenced by factors such as age and genetic polymorphisms rather than smoking,^46,47^ CAA-related pathology may have masked the potential effects of smoking on ICH risk in the CMB(+) group. This does not necessarily mean that smoking has no impact on ICH in patients with CMBs, but rather suggests that the contribution of CAA, a pathology independent of smoking exposure, may make it difficult to detect the effects of smoking in this population. While the analogy of masking by other pathologies, such as CAA, to explain why smoking was not associated with ICH in the CMB(+) group is plausible, it remains speculative. Further studies focusing on CAA-specific cohorts are necessary to delineate this relationship more clearly.

In contrast, supporting our findings in the CMB(–) group, a previous meta-analysis suggested that smoking tends to be associated with non-lobar ICH (pooled risk ratio: 1.24, 95% CI: 0.98–1.58).^10^ This tendency might reflect the greater impact of smoking on hypertensive vasculopathy rather than CAA-related pathology, consistent with our finding of significant associations only in the CMB(–) group. Furthermore, while our age-stratified analysis showed no association between current smoking and ICH, our hypertension-stratified analysis revealed significantly elevated ICH risk in individuals with hypertension without CMBs. This differential influence of smoking based on CMB status may explain the inconsistent findings in previous studies on smoking-ICH associations,^6,7,10,11^ where CMB status was not considered as an effect modifier.

In the present study, long-term smoking cessation was significantly associated with decreased odds of ICH compared with current smoking, particularly in the CMB(–) group. A previous study comparing current to past smokers did not observe a significant risk reduction^12^; however, it did not stratify individuals on the basis of CMB status. In SAH, smoking cessation effectively prevents aneurysm rupture.^48^ Our findings demonstrated similar preventive benefits against ICH in the CMB(–) group.

This study has several strengths. First, we included a substantial sample size of 487 patients with ICH and 322 controls, which provided adequate statistical power for our analyses. Second, CMBs were evaluated using standardized assessment protocols according to the STRIVE criteria on GRE T2*WI. This approach ensured consistent and reliable identification across all participants. Third, our approach of stratifying by CMB status clarified the association between ICH and smoking and provided insights into the heterogeneity of ICH pathophysiology.

This study also has some limitations. First, the retrospective, single-center design conducted in a Japanese institution may have introduced selection bias and limited the generalizability of our findings to broader populations. Moreover, our participant selection process, with major exclusions due to missing CMB evaluation and smoking status, as we focused on first-ever ICH events to better understand how ICH develops in individuals without previous cerebrovascular events, may have systematically affected the representativeness of our study population and potentially led to either overestimation or underestimation of the association between ICH and smoking. Second, smoking status was self-reported, which may have led to recall bias and underreporting of smoking habits, especially among past smokers. The categorization of smoking cessation into ≤10 or >10 years, while practical, is somewhat arbitrary and may not fully capture nuanced risk reductions over time. These measurement limitations could have resulted in misclassification bias, potentially attenuating the observed associations. Third, while we adjusted for major risk factors, residual confounding from unmeasured variables, such as secondhand smoke exposure, environmental factors, and socioeconomic factors (e.g., education, income), cannot be excluded. Fourth, the controls were selected from among patients without stroke but with neurological diseases. This lack of healthy controls could introduce selection bias, as these individuals may have other risk factors not accounted for in the analysis. This is exemplified in the Control/CMB(+) group, which showed higher antithrombotic use and higher premorbid mRS, despite having no history of stroke. This selection bias could have reduced our ability to detect true smoking effects. Finally, the small population of the Control/CMB(+) group, due to our selection, potentially limited the statistical power of our analyses. Additionally, missing data were handled using pairwise deletion, which may introduce bias. These limitations are reflected in the wide confidence intervals observed in some subgroups, such as short-term cessation in the CMB(+) group, indicating low statistical power. This limitation should be considered when interpreting the results, as it may obscure potential associations. These methodological constraints necessitate a cautious interpretation of our subgroup findings, which should be considered exploratory rather than definitive.

In conclusion, this study demonstrated a significant association between current smoking status and increased ICH risk, particularly in individuals without CMBs, where hypertensive vasculopathy predominates. Additionally, long-term smoking cessation was associated with a substantial decrease in ICH risk. Clinically, recognizing smoking as a modifiable risk factor for ICH in individuals without CMBs provides an opportunity to reduce its incidence and improve outcomes. Assessment of smoking history should be incorporated into ICH risk evaluation, and early smoking cessation should be emphasized as a preventive strategy in this population. Future research should address these limitations by incorporating prospective designs, objective measures of smoking exposure, multicenter data, and healthy controls to improve the external validity across diverse populations and explore the underlying mechanisms.

## Data Availability

Data supporting the findings of this study are available from the corresponding author on reasonable request.

## Acknowledgments

We would like to express our sincere gratitude to the patients who contributed their data to this study, as well as to all the individuals involved in data collection and management.

## Supplemental Material

Tables S1–S4

## Non-standard Abbreviations and Acronyms

aOR: adjusted odds ratio
AIS: acute ischemic stroke
BBB: blood-brain barrier
BMI: body mass index
CAA: cerebral amyloid angiopathy
CBA: Charcot–Bouchard aneurysm
CI: confidence interval
CMB: cerebral microbleed
GRE T2*WI: gradient-echo T2 star-weighted imaging
HR: hazard ratio
ICH: intracerebral hemorrhage
IQR: interquartile range
MRI: magnetic resonance imaging
mRS: modified Rankin Scale
NCVC: National Cerebral and Cardiovascular Center
OR: odds ratio
ROS: reactive oxygen species
SAH: subarachnoid hemorrhage
STRIVE: Standards for Reporting Vascular Changes on Neuroimaging
TNF-α: tumor necrosis factor-alpha

